# Rapid increase in Omicron infections in England during December 2021: REACT-1 study

**DOI:** 10.1101/2021.12.22.21268252

**Authors:** Paul Elliott, Barbara Bodinier, Oliver Eales, Haowei Wang, David Haw, Joshua Elliott, Matthew Whitaker, Jakob Jonnerby, David Tang, Caroline E. Walters, Christina Atchison, Peter J. Diggle, Andrew J. Page, Alexander J. Trotter, Deborah Ashby, Wendy Barclay, Graham Taylor, Helen Ward, Ara Darzi, Graham S. Cooke, Marc Chadeau-Hyam, Christl A. Donnelly

## Abstract

**Background:** The highest-ever recorded numbers of daily severe acute respiratory syndrome coronavirus 2 (SARS-CoV-2) infections in England has been observed during December 2021 and have coincided with a rapid rise in the highly transmissible Omicron variant despite high levels of vaccination in the population. Although additional COVID-19 measures have been introduced in England and internationally to contain the epidemic, there remains uncertainty about the spread and severity of Omicron infections among the general population.

**Methods:** The REal-time Assessment of Community Transmission–1 (REACT-1) study has been monitoring the prevalence of SARS-CoV-2 infection in England since May 2020. REACT-1 obtains self-administered throat and nose swabs from a random sample of the population of England at ages 5 years and over. Swabs are tested for SARS-CoV-2 infection by reverse transcription polymerase chain reaction (RT-PCR) and samples testing positive are sent for viral genome sequencing. To date 16 rounds have been completed, each including ∼100,000 or more participants with data collected over a period of 2 to 3 weeks per month. Socio-demographic, lifestyle and clinical information (including previous history of COVID-19 and symptoms prior to swabbing) is collected by online or telephone questionnaire. Here we report results from round 14 (9-27 September 2021), round 15 (19 October - 05 November 2021) and round 16 (23 November - 14 December 2021) for a total of 297,728 participants with a valid RT-PCR test result, of whom 259,225 (87.1%) consented for linkage to their NHS records including detailed information on vaccination (vaccination status, date). We used these data to estimate community prevalence and trends by age and region, to evaluate vaccine effectiveness against infection in children ages 12 to 17 years, and effect of a third (booster) dose in adults, and to monitor the emergence of the Omicron variant in England.

**Results:** We observed a high overall prevalence of 1.41% (1.33%, 1.51%) in the community during round 16. We found strong evidence of an increase in prevalence during round 16 with an estimated reproduction number R of 1.13 (1.06, 1.09) for the whole of round 16 and 1.27 (1.14, 1.40) when restricting to observations from 1 December onwards. The reproduction number in those aged 18-54 years was estimated at 1.23 (1.14, 1.33) for the whole of round 16 and 1.41 (1.23, 1.61) from 1 December. Our data also provide strong evidence of a steep increase in prevalence in London with an estimated R of 1.62 (1.34, 1.93) from 1 December onwards and a daily prevalence reaching 6.07% (4.06%, 9.00%) on 14 December 2021. As of 1 to 11 December 2021, of the 275 lineages determined, 11 (4.0%) corresponded to the Omicron variant. The first Omicron infection was detected in London on 3 December, and subsequent infections mostly appeared in the South of England. The 11 Omicron cases were all aged 18 to 54 years, double-vaccinated (reflecting the large numbers of people who have received two doses of vaccine in this age group) but not boosted, 9 were men, 5 lived in London and 7 were symptomatic (5 with classic COVID-19 symptoms: loss or change of sense of smell or taste, fever, persistent cough), 2 were asymptomatic, and symptoms were unknown for 2 cases. The proportion of Omicron (vs Delta or Delta sub-lineages) was found to increase rapidly with a daily increase of 66.0% (32.7%, 127.3%) in the odds of Omicron (vs. Delta) infection, conditional on swab positivity. Highest prevalence of swab positivity by age was observed in (unvaccinated) children aged 5 to 11 years (4.74% [4.15%, 5.40%]) similar to the prevalence observed at these ages in round 15. In contrast, prevalence in children aged 12 to 17 years more than halved from 5.35% (4.78%, 5.99%) in round 15 to 2.31% (1.91%, 2.80%) in round 16. As of 14 December 2021, 76.6% children at ages 12 to 17 years had received at least one vaccine dose; we estimated that vaccine effectiveness against infection was 57.9% (44.1%, 68.3%) in this age group. In addition, the prevalence of swab positivity in adults aged 65 years and over fell by over 40% from 0.84% (0.72%, 0.99%) in round 15 to 0.48% (0.39%,0.59%) in round 16 and for those aged 75 years and over it fell by two-thirds from 0.63% (0.48%,0.82%) to 0.21% (0.13%,0.32%). At these ages a high proportion of participants (>90%) had received a third vaccine dose; we estimated that adults having received a third vaccine dose had a three- to four-fold lower risk of testing positive compared to those who had received two doses.

**Conclusion:** A large fall in swab positivity from round 15 to round 16 among 12 to 17 year olds, most of whom have been vaccinated, contrasts with the continuing high prevalence among 5 to 11 year olds who have largely not been vaccinated. Likewise there were large falls in swab positivity among people aged 65 years and over, the vast majority of whom have had a third (booster) vaccine dose; these results reinforce the importance of the vaccine and booster campaign. However, the rapidly increasing prevalence of SARS-CoV-2 infections in England during December 2021, coincident with the rapid rise of Omicron infections, may lead to renewed pressure on health services. Additional measures beyond vaccination may be needed to control the current wave of infections and prevent health services (in England and other countries) from being overwhelmed.

**Summary:** The unprecedented rise in SARS-CoV-2 infections is concurrent with rapid spread of the Omicron variant in England and globally. We analysed prevalence of SARS-CoV-2 and its dynamics in England from end of November to mid-December 2021 among almost 100,000 participants from the REACT-1 study. Prevalence was high during December 2021 with rapid growth nationally and in London, and of the proportion of infections due to Omicron. We observed a large fall in swab positivity among mostly vaccinated older children (12-17 years) compared with unvaccinated younger children (5-11 years), and in adults who received a third vs. two doses of vaccine. Our results reiterate the importance of vaccination and booster campaigns; however, additional measures may be needed to control the rapid growth of the Omicron variant.

## Introduction

In many settings, vaccination against the SARS-CoV-2 virus has been shown to have reduced COVID-19-related hospital admissions and deaths [1,2]. It has been estimated that vaccinations prevented 7.2 million infections and 27,000 deaths in England alone up to June 2021[3] and could have prevented 163,000 deaths in the US between June and December 2021 [4]. However, the appearance of SARS-CoV-2 variants with increased transmissibility is an ongoing threat around the world [5,6]. The recent emergence of the Omicron variant has led to travel restrictions, originally for those travelling from southern Africa, but recently for those wishing to travel from the UK. The Omicron variant has spread rapidly both in South Africa and in regions of the UK, driven by the ability of Omicron to cause more breakthrough infections among vaccinated individuals than other variants, likely due to the large number of genetic mutations within the viral spike protein [7]. Furthermore, vaccine-induced protection against the Delta variant (and its sub-lineages) was already found to be waning [8].

The REal-time Assessment of Community Transmission-1 (REACT-1) study [9–11] has been documenting the spread of the SARS-CoV-2 virus in England approximately monthly since May 2020 as the first wave of infections in England declined. REACT-1 charted the complete replacement of the Alpha variant by the Delta variant from round 12 (21 May to 7 June 2021) to round 13 (24 June to 12 July 2021) [11]. With round 16 (23 November to 14 December 2021) we are similarly able to document Omicron’s early spread in England.

In response to the pandemic, the national vaccination programme in England is evolving quickly. In September 2021 in addition to the vaccinations offered to those 16 years of age and over, school-aged children aged 12 to 15 years were offered a single dose of vaccine, and third (booster) doses were made available to health and social care workers, all those aged 50 years and over as well as younger people at risk. By December 2021, the opportunity to schedule booster doses has been extended to all adults (18 years of age and older) with heightened efforts to deliver the booster doses as quickly as possible. At the same time, there has been an accelerated rollout of the vaccination programme to older school-aged children (12 to 17 years) with double-doses now being offered to 12 to 15 year-olds as well as 16 and 17 year-olds [12].

Here we document the early detection of Omicron variant in England using the community-based REACT-1 study to avoid the biases that arise in case incidence data, including those arising to due test-seeking behaviour and limits on testing capacity [10]. We compare SARS-CoV-2 swab positivity in round 16 (23 November to 14 December 2021) to previous rounds and analyse vaccine effectiveness against infection in school-aged children aged 12 to 17 years and the effect of the third (booster) dose in adults (compared to those with with two doses) via linkage to National Health Service (NHS) vaccination data for consenting participants.

## Results

In the sixteenth round of REACT-1, 803,864 randomly selected individuals aged 5 years and over in England were invited to take part in the study, with data collection from 23 November to 14 December 2021. Of those invited, 129,534 (16.1%) registered of whom 97,089 (75.0%) provided a self-administered throat and nasal swab with a valid RT-PCR test result, including 661 samples (12 positives) obtained from 15 to 17 December 2021 (see **Materials and Methods**). A total of 1,192 positive swabs were detected yielding an overall weighted prevalence of 1.41% (1.33%, 1.51%), which represents the third highest prevalence observed since the beginning of REACT-1 (1 May 2020) (Supplementary Table 1).

### Temporal and geographical trends

Fitting a P-spline regression model to all REACT-1 data, we found clear evidence for an increase in the weighted prevalence during round 16 starting around 1 December 2021 (Figure 1 A). We estimated an overall reproduction number R=1.13 (1.06, 1.19) for the whole of round 16, based on an exponential growth/decay model for the daily weighted prevalence. Restricting to data from 1 December onwards, the estimate of R was 1.27 (1.14, 1.40), with posterior probability that R>1 of >0.99 in both cases (Table 1). The exponential growth/decay model fit to round 16 data indicated an increase in weighted prevalence in those aged 18 to 54 years with R=1.23 (1.14, 1.33) for the whole of round 16 and R=1.41 (1.23, 1.61) for data restricted to 1 December onwards, both with posterior probability that R>1 of >0.99 (Table 1). Flexible P-spline models for three broad age groups also indicated an increasing prevalence in those aged 18 to 54 years (Figure 1 B).

**Table 1.** Table of growth rates, reproduction numbers and doubling/halving times from exponential model fits on data from round 16 (23 November to 14 December 2021)^1^

| Rounds |  |  | Growth rate | R | Probability R>1 | Doubling (+) / Halving (-) time* |
| --- | --- | --- | --- | --- | --- | --- |
| 16 | All positives |  | 0.019 ( 0.010 , 0.029 ) | 1.13 ( 1.06 , 1.19 ) | >0.99 | 36.1 ( 71.7 , 24.1 ) |
|  | Age | Aged 17 and under | -0.019 ( -0.035 , -0.004 ) | 0.88 ( 0.79 , 0.98 ) | 0.01 | -35.7 ( -19.6 , ** ) |
|  |  | Aged 18 to 54 | 0.034 ( 0.021 , 0.047 ) | 1.23 ( 1.14 , 1.33 ) | >0.99 | 20.3 ( 33.6 , 14.6 ) |
|  |  | Aged 55 and over | 0.005 ( -0.022 , 0.032 ) | 1.03 ( 0.86 , 1.21 ) | 0.64 |  |
|  | Region | East Midlands | 0.010 ( -0.020 , 0.040 ) | 1.07 ( 0.88 , 1.27 ) | 0.75 |  |
|  |  | West Midlands | 0.019 ( -0.017 , 0.054 ) | 1.13 ( 0.90 , 1.38 ) | 0.86 |  |
|  |  | East of England | 0.013 ( -0.019 , 0.043 ) | 1.08 ( 0.89 , 1.30 ) | 0.79 |  |
|  |  | London | 0.060 ( 0.040 , 0.080 ) | 1.42 ( 1.27 , 1.58 ) | >0.99 | 11.6 ( 17.5 , 8.7 ) |
|  |  | North West | 0.001 ( -0.032 , 0.034 ) | 1.01 ( 0.81 , 1.23 ) | 0.52 |  |
|  |  | North East | -0.018 ( -0.078 , 0.038 ) | 0.89 ( 0.57 , 1.26 ) | 0.27 |  |
|  |  | South East | 0.022 ( 0.000 , 0.044 ) | 1.15 ( 1.00 , 1.30 ) | 0.97 |  |
|  |  | South West | -0.005 ( -0.035 , 0.024 ) | 0.97 ( 0.79 , 1.16 ) | 0.38 |  |
|  |  | Yorkshire and The Humber | -0.022 ( -0.056 , 0.012 ) | 0.87 ( 0.68 , 1.08 ) | 0.10 |  |
| 16 (December only) | All positives |  | 0.039 ( 0.021 , 0.057 ) | 1.27 ( 1.14 , 1.40 ) | >0.99 | 17.8 ( 32.9 , 12.2 ) |
|  | Age | Aged 17 and under | -0.014 ( -0.045 , 0.017 ) | 0.91 ( 0.74 , 1.11 ) | 0.19 |  |
|  |  | Aged 18 to 54 | 0.058 ( 0.034 , 0.083 ) | 1.41 ( 1.23 , 1.61 ) | >0.99 | 11.9 ( 20.2 , 8.4 ) |
|  |  | Aged 55 and over | 0.011 ( -0.044 , 0.063 ) | 1.07 ( 0.74 , 1.45 ) | 0.65 |  |
|  | Region | East Midlands | 0.047 ( -0.009 , 0.101 ) | 1.32 ( 0.95 , 1.77 ) | 0.95 |  |
|  |  | West Midlands | -0.018 ( -0.084 , 0.045 ) | 0.89 ( 0.54 , 1.31 ) | 0.29 |  |
|  |  | East of England | 0.032 ( -0.028 , 0.090 ) | 1.22 ( 0.83 , 1.67 ) | 0.86 |  |
|  |  | London | 0.085 ( 0.050 , 0.120 ) | 1.62 ( 1.34 , 1.93 ) | >0.99 | 8.2 ( 14.0 , 5.8 ) |
|  |  | North West | -0.004 ( -0.070 , 0.058 ) | 0.98 ( 0.61 , 1.41 ) | 0.46 |  |
|  |  | North East | 0.067 ( -0.051 , 0.186 ) | 1.48 ( 0.71 , 2.59 ) | 0.87 |  |
|  |  | South East | 0.047 ( 0.007 , 0.087 ) | 1.33 ( 1.04 , 1.64 ) | 0.99 | 14.6 ( ** , 7.9 ) |
|  |  | South West | -0.027 ( -0.089 , 0.032 ) | 0.84 ( 0.52 , 1.22 ) | 0.19 |  |
|  |  | Yorkshire and The Humber | -0.004 ( -0.072 , 0.062 ) | 0.98 ( 0.60 , 1.44 ) | 0.46 |  |

\* Doubling/Halving time estimates are shown only when the 95% credible intervals for R exclude 1
\*\* Doubling/Halving time had an estimated magnitude greater than 50 days and so represented approximately constant prevalence
<sup>1</sup> Includes N=661 samples (12 positives) obtained from 15-17 December 2021

**Figure 1.**
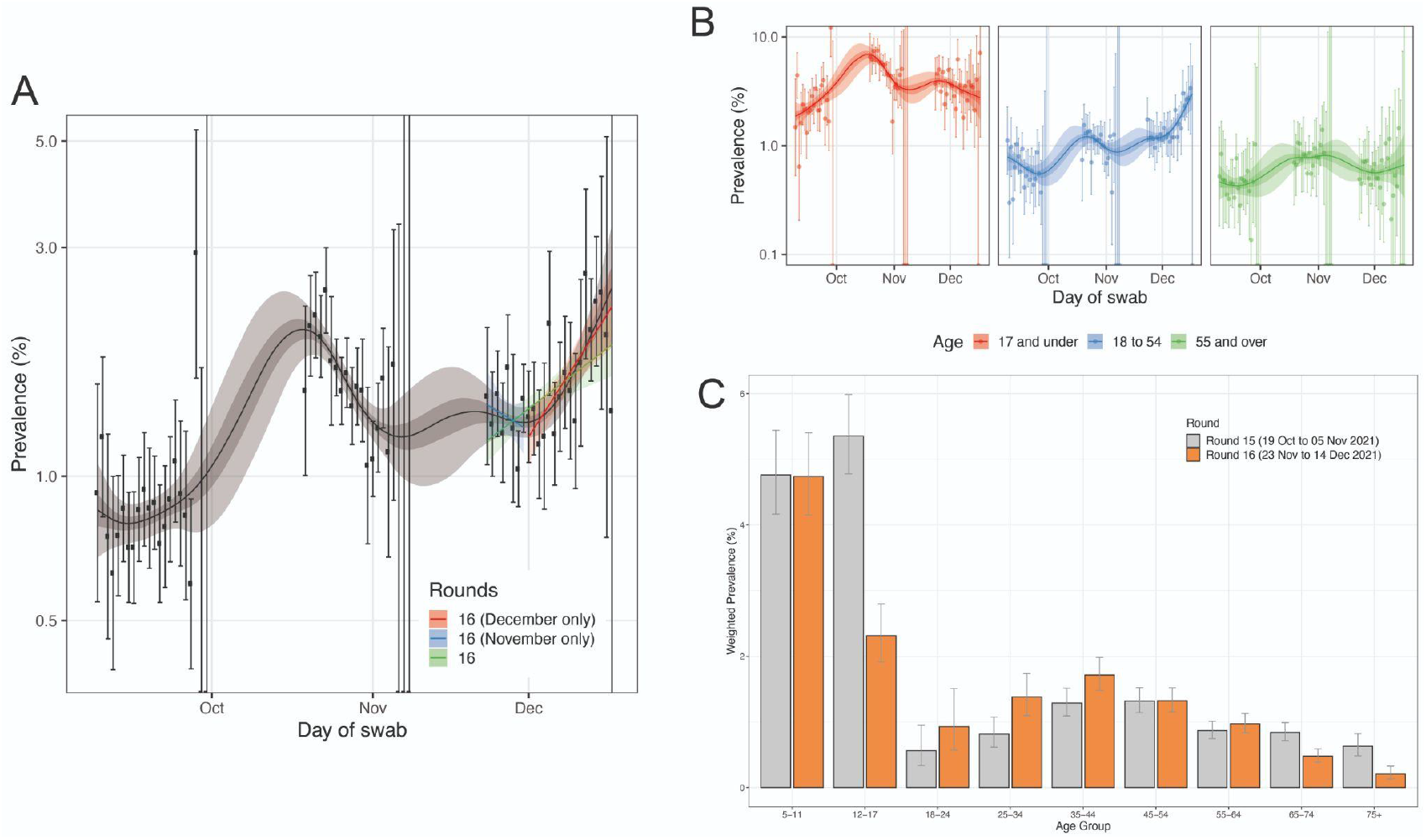
**(A)** Comparison of exponential model fits to round 16 (green), round 16 from 23-30 November (blue) and from 1 December onwards (red) in addition to a P-spline model fit to all rounds of REACT-1 (black, shown here only for rounds 14, 15 and 16). Shaded blue and red regions show the 95% posterior credible interval for the exponential models, and the shaded grey region shows 50% (dark grey) and 95% (light grey) posterior credible interval for the P-spline model. Results are presented for each day (X axis) of sampling for round 14, round 15 and round 16 and the prevalence of swab positivity is shown (Y axis) on a log scale. Weighted observations (black dots) and 95% confidence intervals (vertical lines) are also shown. **(B)** P-spline models fit to all rounds of REACT-1 for those aged 17 years and under (red), those aged 18 to 54 years inclusive (blue) and those aged 55 years and over (green). **(C)** Weighted prevalence of swab positivity by age group for round 15 and round 16. Bars show the prevalence point estimates (grey for round 15 and orange for round 16), and the vertical lines represent the 95% confidence intervals.

We found strong evidence of high and increasing weighted prevalence in London which had the highest weighted prevalence nationally at 1.84% (1.59%, 2.12%) compared with 1.23% (1.03%, 1.47%) in round 15 (Figure 2 A, Supplementary Table 4). Using the exponential growth/decay model fit to daily weighted prevalence in London, we estimated an R of 1.42 (1.27, 1.58) in round 16 and 1.62 (1.34, 1.93) from 1 December 2021, both with >0.99 posterior probability that R>1 (Table 1).

**Figure 2.**
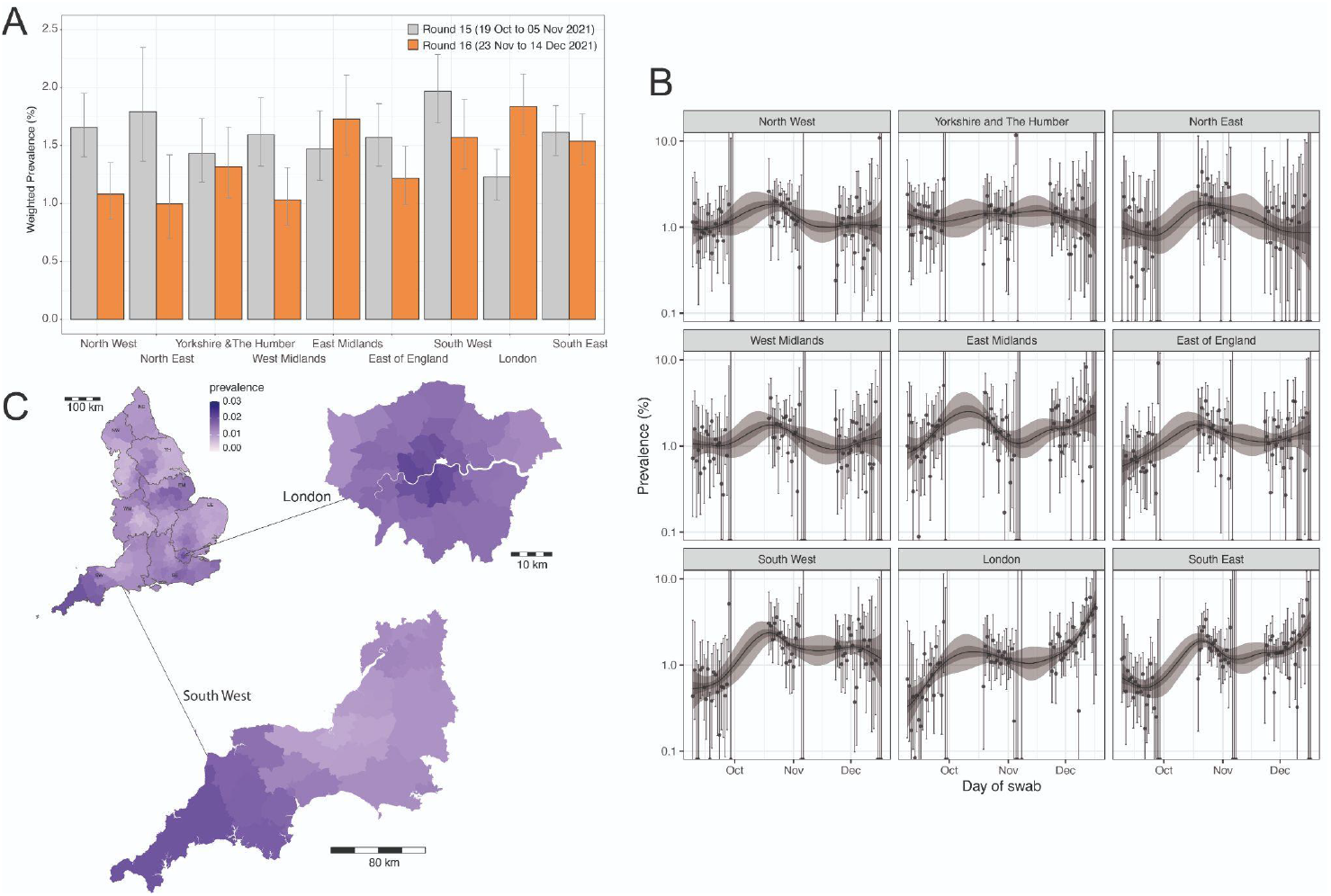
**(A)** Weighted prevalence of swab positivity by region for round 15 and round 16. Bars represent prevalence point estimates (grey for round 15 and orange for round 16), and the vertical lines the corresponding 95% confidence intervals. **(B)** P-spline models fit to all rounds of REACT-1 for each of the nine regions separately. Shown here only for the period of round 14, round 15 and round 16. Shaded regions show 50% (dark shade) and 95% (light shade) posterior credible interval for the P-spline models. **(C)** Neighbourhood smoothed average prevalence by lower-tier local authority area for round 16. Neighbourhood prevalence calculated from nearest neighbours (the median number of neighbours within 30 km in the study). Average neighbourhood prevalence displayed for individual lower-tier local authorities for the whole of England and for South East, South West and London. Regions: NE = North East, NW = North West, YH = Yorkshire and The Humber, EM = East Midlands, WM = West Midlands, EE = East of England, L = London, SE = South East, SW = South West

Daily estimates of weighted prevalence in each region separately show a steep increase in London from 0.80% (0.36%, 1.75%) on 26 November to 6.06% (4.06%, 9.00%) on 14 December (Figure 2 B). A (somewhat slower) increase was also observed in daily weighted prevalence in the South East, which reached 5.75% (2.60%, 12.22%) by 15 December 2021. Using a nearest neighbour approach we estimated smoothed prevalence at the Lower-Tier Local Authority (LTLA) level. Eight of the ten highest smoothed estimates of prevalence over the whole of round 16 were found in London (Lambeth, Kensington and Chelsea, Hammersmith and Fulham, Southwark, Islington, Westminster, Wandsworth, Camden) with smoothed prevalence estimates ranging from 2.15% to 1.94% and the remaining two were in parts of South West (Cornwall, Plymouth) with smoothed prevalence estimates of 1.94% and 1.81%, respectively (Figure 2 C).

### Delta and Omicron infections

Of the 1,048 positive swabs collected up to and including 11 December 2021, 650 lineages were determined with at least 50% genome coverage (Figure 3 A-D, Supplementary Table 2). Of these, 275 were collected from 1 to 11 December, 2021, of which 11 (4.0%) were Omicron variants, all others being Delta or Delta sub-lineages. The first swab testing positive for Omicron variant was obtained on 3 December 2021 (Supplementary Table 3) in London (Figure 3 B). Subsequent infections were all detected in the South of England except for one in East Midlands (Figure 3 C-D). The proportion of Omicron (vs Delta or Delta sub-lineages) appears to have rapidly increased (Figure 3 E), with a daily increase of 66.0% (32.7%, 127.3%) in the odds of Omicron (vs. Delta) infection, conditional on testing positive. Assuming the dynamics of Omicron growth (and Delta’s) remain constant, it is estimated to take 8.7 (5.4, 15.5) days for Omicron to increase from a proportion of 10% to 90%. This is approximately 3.5 times faster than the estimated 31.4 (43.9, 22.0) days it took for Delta to grow from 10% to 90% against Alpha. Assuming 100% sensitivity and a weighted prevalence of 1.38% from 1 to 11 December 2021, during which the 11 Omicron variants were detected (out of 275 positives sequenced), we estimated a prevalence of Omicron infections in England of 31,000 (17,000, 55,000) averaged over that period, but at a time that prevalence was increasing rapidly.

**Figure 3.**
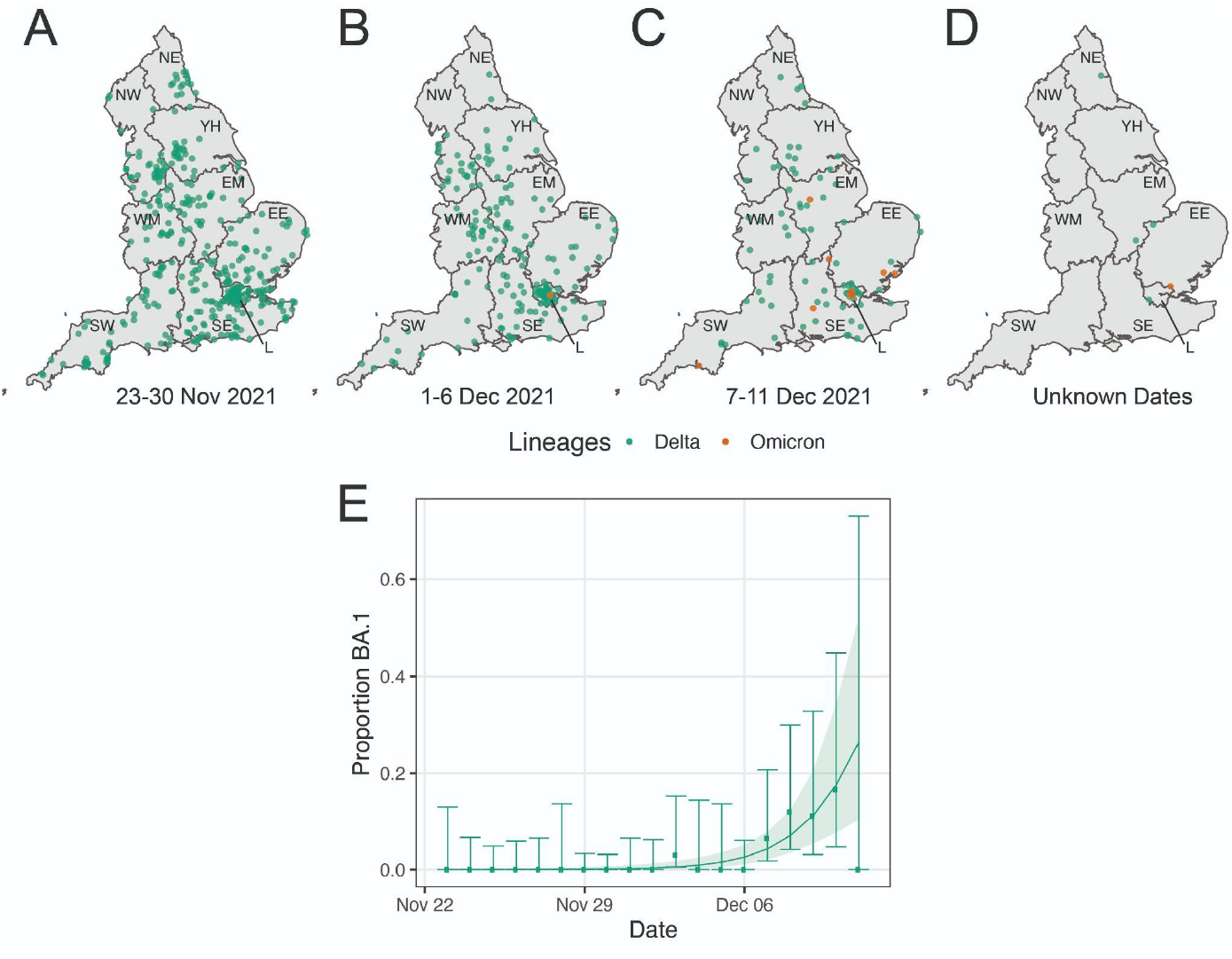
Geographical distribution (jittered) of the (N=650) positive swabs with determined lineages and at least 50% genome coverage. Delta infections are presented in green, and Omicron infections in orange. Results are presented for the (N=378) infections detected from 23 November to 30 November 2021 **(A)**, the (N=188) infections detected from 1 December to 6 December 2021 **(B)**, the (N=87) infections detected from 7 December to 11 December 2021 **(C)** and for the (N=5) infections with unknown dates of swabbing **(D)**. Regions: NE = North East, NW = North West, YH = Yorkshire and The Humber, EM = East Midlands, WM = West Midlands, EE = East of England, L = London, SE = South East, SW = South West. Daily proportion of Omicron infections among positive swabs with determined lineage and at least 50% genome coverage in round 16. Point estimates are represented (dots) along with 95% confidence intervals (vertical lines). Smoothed estimates of the proportion are also shown (solid line) together with their 95% credible intervals (shaded regions) **(E)**.

The 11 Omicron-infected participants were all aged 18 to 54 years, double-vaccinated (reflecting the large numbers of people who have received two doses of vaccine in this age group) and were not boosted, 9 (81.8%) were men, 5 (45.4%) lived in London and 7 (63.6%) were symptomatic: 5 (45.4%) reported classic COVID-19 symptoms (loss or change of sense of taste or smell, fever, persistent cough), 2 (18.2%) were asymptomatic, and symptoms were unknown for 2 cases.

### Variation in prevalence by age

The highest weighted prevalence in round 16 by age was observed in those aged 5 to 11 years at 4.74% (4.15%, 5.40%) which was similar to that observed in round 15 at 4.76% (4.16%, 5.44%) (Figure 1 C, Supplementary Table 4). In contrast, between rounds 15 and 16 weighted prevalence fell from 5.35% (4.78%, 5.99%) to 2.31% (1.91%, 2.80%) in those aged 12 to 17 years, from 0.84% (0.72%, 0.99%) to 0.48% (0.39%, 0.59%) in those aged 65 to 74 years, and from 0.63% (0.48%, 0.82%) to 0.21% (0.13%, 0.32%) in those aged 75 years and over.

### Effects of vaccine on swab positivity

Using data from rounds 14, 15 and 16 (September to December 2021), we estimated an adjusted vaccine effectiveness against infection in children aged 12 to 17 years of 55.7% (41.3%, 66.6%) for those having received a single vaccine dose and of 57.9% (44.1%, 68.3%) for those having received one or two vaccine doses (Supplementary Table 5). Over the whole period of round 16 (27 November, 17 December 2021) 76.6% of the participants aged 12 to 17 years had received one or two vaccine doses more than 14 days prior to swabbing (Supplementary Figure 1). Our estimates of vaccine effectiveness against infection together with the contrast between the continuing high prevalence in (unvaccinated) children aged 5 to 11 years and the fall in prevalence in the 12 to 17 year-olds, are indicative of the impact of the vaccination programme in children.

Our data also demonstrate the contribution of a third (booster) vaccine dose to the fall in prevalence we observed in round 16 in those aged 65 years and over. Over round 16, 91.2% and 96.8% of participants aged 65 to 74 years and 75 years and over, respectively, had received a third vaccine dose more than 14 days prior to swabbing (Supplementary Figure 1). We estimated an adjusted odds ratio (OR) for swab positivity of 0.27 (0.22, 0.34) for adults aged 18 years and over who had received three vaccine doses compared to those having received two doses (Supplementary Table 6).

### Socio-demographic trends

We found highest weighted prevalence in round 16 in larger households including 5 people at 2.73% (2.25%, 3.32%) and 6 or more people at 2.65% (2.00%, 3.50%) compared to 0.88% (0.72%, 1.09%) in single-person households; in households with one or more children at 2.43% (2.23%, 2.65%) compared to 0.85% (0.76%, 0.95%) in households without children; in those having been in contact with a confirmed COVID-19 case at 8.00% (7.25%, 8.82%) compared to 0.81% (0.73%, 0.89%) for those without such contact, and in those reporting classic COVID-19 symptoms in the month prior to swabbing at 6.96% (6.32%, 7.67%) compared to 0.62% (0.55%, 0.70%) in those without symptoms (Supplementary Table 7). Multivariable logistic regression (Supplementary Table 8) showed increased risk of swab positivity for those living with one or more children in the household (mutually adjusted OR 1.37 (1.12, 1.67) compared to households with no children) and for those living in households of 6 or more people (mutually adjusted OR=1.39 (1.01, 1.91) compared to single-person households), suggesting high levels of within-household transmission. Risk of swab positivity was also higher in participants living in areas in the second most compared to the least deprived quintile of the Index of Multiple Deprivation (IMD)[13] with mutually adjusted OR of 1.28 (1.07, 1.52), and in other essential workers with mutually adjusted OR of 1.18 (1.01, 1.38) compared to other workers. In contrast, our results suggest that health care and care home workers were at lower risk of swab positivity with mutually adjusted OR of 0.75 (0.59, 0.94) compared to other workers.

## Discussion

The sixteenth round of the REACT-1 study (23 November to 14 December 2021) followed a continuous period of high prevalence of SARS-CoV-2 in England from summer 2021 [11,14], dominated by the Delta variant until early-mid December 2021. However, since the first reports of the spread of the Omicron variant emerged in South Africa in late November 2021[15], countries around the world have seen a rapid rise in COVID-19 attributed to the Omicron variant [16]. S-gene target failure (SGTF) on RT-PCR testing of samples processed at TaqPath laboratories can be used as a proxy to monitor the spread of Omicron versus Delta [17]. In England, the first Omicron infection was recorded on 27 November and since then it has become the dominant variant, with 71.5% of samples having SGTF of those dated 18-19 December 2021 and processed via TaqPath laboratories[18]. Based on these data, estimates of the regional doubling time for Omicron in England range from 1.6 to 2.5 days; London has the highest regional proportion of SGTF at 89.7%, followed by the East of England at 76.6% [18]. At the same time, the vaccine rollout in England has been accelerating to include a single dose vaccine in children aged 12 to 15 years (double-dose from 20 December), double-dose at ages 16 to 17 years, and a booster programme which since 12 December is being offered to all adults in England.

Against this backdrop we observed a mixed picture in round 16, characterised by i) falling prevalence of swab positivity in older school-aged children (12 to 17 years) where high levels of single- or double-dose vaccination has been taking place, ii) high and constant prevalence in younger school-aged children (5 to 11 years) who are largely ineligible for vaccination, iii) falling prevalence among older people (65 years and over) who have largely had booster vaccinations, and iv) since around 1 December 2021, rapidly rising prevalence nationally and especially in London and the South of England, coincident with the rapid rise of Omicron variant.

In our own data we detected 11 Omicron infections in swabs obtained up to 11 December 2021, as the Omicron epidemic took off in England. This coincided with a rapidly rising proportion of Omicron compared to Delta infections detected via sequencing, which likely reflects both the rapid growth of Omicron and the replacement of Delta by Omicron [18]. Household transmission from an Omicron index case is reported to be approximately three times higher than that of Delta [6], which may at least in part explain its transmission advantage. It has been suggested that Omicron may be associated with less severe disease than Delta with the caveat of differential population immunity from vaccination and previous infection [19]. However, the small number of Omicron infections observed in REACT-1 to date were among young to middle-aged adults where case fatality ratios will be lower than among the older population.

Omicron may have greater escape from immunity conferred by vaccination than Delta, with an estimated 20 to 40 times higher antibody titre required for neutralisation [20]. *In vitro* studies show clear evidence that individuals receiving a third (booster) dose of mRNA vaccine following two previous vaccine doses have greatly increased neutralisation of the Omicron variant [7,21]. Although in our study, none of the confirmed Omicron infections were observed in individuals following booster immunisation (all had received two vaccine doses), the number of Omicron positive samples available at the time of publication is too small to draw conclusions about the effectiveness of boosters against Omicron infections.

While we estimated there were on average 31,000 infections in England from Omicron during 1-11 December, the rapid doubling time suggests that without further measures this figure could reach into the hundreds of thousands by late December 2021. The most recent estimates from the UK Health Security Agency (UKHSA) indicate that there were 60,508 confirmed Omicron cases in England, and 143,252 cumulative cases to 20 December 2021 based on SGTF [18]. However, these figures are likely underestimates, since they depend on people presenting for testing, and unlike community-based studies such as REACT-1, they do not include asymptomatic cases. As testing capacity becomes saturated by rising demand, there will be even greater need for estimates from community surveillance studies and a greater focus on hospitalisations as the epidemic progresses.

Our findings in older school-aged children (12 years and over) amongst whom the vaccine programme (one and two doses) has been accelerated are encouraging. Since round 15 (19 October to 5 November 2021) we saw prevalence in this group drop by over a half, while it remained more or less unchanged amongst primary school-aged children (5 to 11 years). This strongly suggests that the vaccine programme, even a single dose, is effective at these ages. Accordingly, we report here estimates of vaccine effectiveness against infection (assumed to be mainly Delta) of 55.7% for a single dose and 57.9% for one or two doses of vaccine. Our findings are similarly encouraging among older people (ages 65 years and over), most of whom have received a booster dose. In these age groups, prevalence from round 15 fell by over 40% at ages 65 to 74 years and by two-thirds among those aged 75 years and over.

Our study has limitations. Our response rate was 12.1% (i.e. returned and valid tests compared with number of invitations), which, despite correction of our prevalence estimates using rim weighting [22], may have introduced bias into our estimates. Our switch to postal from courier returns meant that some samples were obtained and posted after the nominated closing date; these samples were however processed in the usual way and the results included in our estimates. Although all positive samples on RT-PCR were sent for sequencing, reliable sequence data (at least 50% genome coverage) were obtained on around 60% of samples; with Omicron being first detected in our data for a sample collected on 3 December, our ability to monitor the spread of Omicron versus Delta lineages was limited by small numbers. Nonetheless we were able to detect the rapid increase in proportion of sequenced cases due to Omicron up to 11 December.

Our estimates of vaccine effectiveness depend on comparability of the vaccinated and unvaccinated groups in all respects other than vaccination itself. For the school-aged children aged 12 years and over who were eligible for vaccination, the rollout was such that not all children were able to access vaccination at the same time making it more likely that vaccinated and unvaccinated children were similar in other respects. Likewise, in comparing double-vaccinated individuals with those having had a third (booster) dose, the assumption is that the two groups are similar other than their vaccine history. This assumption appears to be valid to the extent that to receive the booster dose, people will need to have been double-vaccinated. This is in contrast to comparisons between double-vaccinated and unvaccinated adults, where, in a highly vaccinated population, characteristics of the two groups are likely to be different [23].

The evidence to suggest that Omicron may cause less severe disease than Delta primarily originates in South Africa [19,24], where the population is comparatively younger than in England and where previous, very severe SARS-CoV-2 waves were driven by wild type, Beta and Delta variants (rather than wild type, Alpha and Delta in England) [25]. Differences between populations and their rates of vaccination and previous infection might limit how much can be extrapolated from other settings such as South Africa to England. One indicative example is that during the week of 12-19 December, COVID-19 hospitalisations rose over 50% in London [26], a pattern not observed in areas with lower prevalence of Omicron. An increasing proportion of the population will have been vaccinated or experienced infection, and therefore may be less likely to have severe disease with Omicron infection compared to those who remain uninfected and not fully vaccinated. Even if infection with Omicron were less severe, the reduced risk of hospitalisation could be rapidly offset by its ongoing exponential growth in England. Close attention therefore needs to be paid to hospitalisation data in England (particularly in London) over the coming weeks, given the lag between infection and hospitalisation.

In summary, we found strong evidence for the effectiveness of vaccination in adolescents and for booster vaccinations against infection in older populations, presumed to be mainly against the Delta variant in our data so far. However, Delta is rapidly being replaced in England by the Omicron variant and further data are needed to confirm the clinical relevance of the strong *in vitro* evidence of booster vaccines protecting against Omicron [7,27]. Moreover, given that booster vaccines may take up to two weeks to have their full protective effect [28], it is likely that a vaccination programme alone may not be sufficient to control the growth of Omicron in the short term. Additional measures beyond vaccination may be needed to control the current wave of infections and prevent health services (in England and other countries) from being overwhelmed.

## Materials and methods

### Study population and sampling

The REACT-1 study methods are published [9]. We have carried out cross-sectional surveys of a random sample of the population of England, aged 5 years and over, over a two- to three-week period each month since May 2020, except for December 2020 and August 2021. Here we report results for round 16 of REACT-1 (23 November to 14 December 2021, N=97,089 including 661 samples [12 positives] 15-17 December) for participants who provided a valid self-administered throat and nose swab for testing for SARS-CoV-2 by reverse transcription polymerase chain reaction (RT-PCR). (A test was recorded as positive if both N gene and E gene targets were detected or if N gene was detected with cycle threshold (Ct) value below 37.) We compare results with those obtained during round 15 (19 October to 5 November 2021, N=100,112 including 93 [all negative] from 6-8 November) and also include data from round 14 (9 to 27 September 2021, N=100,527 including 509 samples from 28-30 September) [14]. Response rates for the three rounds (number of valid swabs returned divided by number of invites sent out) were 12.2%, 11.7% and 12.1% for rounds 14, 15 and 16 respectively. We used the National Health Service (NHS) list of patients registered with a general practitioner in England as sampling frame, based on data held by NHS Digital who provided information on age, sex and residential location. Participants provided additional information on ethnicity, household size, occupation, past medical history, potential contact with a COVID-19 case, symptoms and other variables during registration and through an online or telephone questionnaire [29]. Using the postcode of residence we linked to an area-level Index of Multiple Deprivation (IMD) quintile or decile [13] and urban/rural status based on classification by the Office for National Statistics [30].

From round 1 (1 May to 1 June 2020) to round 11 (15 April to 3 May 2021) we aimed to achieve approximately equal numbers of participants in each lower-tier local authority (LTLA) in England (N=315), but this was altered from round 12 (20 May to 7 June 2021) to invite a random sample in proportion to population size at LTLA level. This increased numbers sampled in densely populated urban areas while reducing the numbers in more rural areas. For both the original and revised sampling methods we used random iterative method (rim) weighting [22] correcting for age, sex, deciles of the IMD, LTLA counts, and ethnic group, to provide estimates that were representative of the population of England as a whole, as previously described [11]. From round 1 to round 13 (24 June to 12 July 2021) we used dry swabs which were collected from the participant’s home by courier and then transported on a cold chain to a single laboratory. However, from round 14 we tested ‘wet’ (saline) swabs sent to the laboratory in a 1:1 randomised experiment either by courier (without cold chain) or by priority post [14]. Since both methods provided similar results, in subsequent rounds (15 and 16) swabs were returned only using the priority postal service. However, the switch from courier to post for return of swabs meant that some samples were obtained after the nominated closing date for the study, to cover any delays in the postal service. Additional swabs received up to the cut-off date for laboratory analysis were included.

### Viral genome sequencing

We sent samples that tested positive to the Quadram Institute, Norwich, UK, for viral genome sequencing using the ARTIC protocol [31] (version 4) for viral RNA amplification and CoronaHiT for preparation of sequencing libraries [32]. We analysed sequencing data using the ARTIC bioinformatic pipeline[31] and assigned lineages using PangoLEARN (version 2021-11-4) [33]. We searched for Omicron defining mutations down to single reads.

### Data analyses

Data analyses were done using R software. We estimated weighted prevalence (and 95% credible intervals) of socio-demographic and other characteristics in round 16 and compared these to estimates from round 15. We then used logistic regression to estimate the effects of these variables on swab positivity with adjustment for age and sex, and also obtained mutually adjusted odds ratios (and 95% confidence intervals) for age, sex, region, employment type, ethnicity, household size, numbers of children in the household, neighborhood deprivation, and urban/rural environment.

We investigated temporal trends in swab positivity using an exponential model of growth or decay with the assumption that numbers of positives out of the total number of samples per day arose from a binomial distribution. We used day of swabbing where reported, otherwise day of first scan of the sample by the Post Office (swabs were excluded from these analyses when neither date was available). We used a bivariate No-U-Turn Sampler with uniform prior for the probability of swab positivity on day of swabbing to estimate posterior credible intervals and the growth rate [34]. We estimated the reproduction number R by assuming a gamma-distributed generation time with shape parameter n=2.29 and rate parameter *β*=0.36 (corresponding to a mean generation time of 6.29 days) [35].

To visualise trends in swab positivity over time, we fit a Bayesian penalised-spline (P-spline) model [36] to the daily data using a No-U-Turn Sampler in logit space. We partitioned the data into approximately 5-day sections by regularly spaced knots, and minimized edge effects by adding further knots beyond the study period. We defined fourth-order basis splines (b-splines) over the knots and guarded against overfitting by including a second-order random-walk prior distribution on the coefficients of the b-splines, with the prior penalising against changes in the growth rate unless supported by the data, as previously described [11]. P-splines were also fit separately to three broad age groups (17 years and under, 18 to 54 years, 55 years and over) and to each region with a smoothing parameter obtained from the model fit to all the data.

We fit a Bayesian logistic regression model to the proportion of lineages identified as the Omicron variant during round 16 to obtain a daily growth rate advantage for Omicron compared with Delta and its sub-lineages.

We estimated geographical variation in prevalence at LTLA level, using a neighborhood spatial smoothing method based on nearest neighbor up to 30 km. We first obtained the median number of study participants in round 16 within 30 km of each study participant, and then calculated the local prevalence for 15 members of each LTLA to estimate the smoothed neighborhood prevalence in that area.

We linked to data from the national COVID-19 vaccination programme to obtain information (with consent) on who had received one or more vaccinations and dates of vaccination. We estimated vaccine effectiveness against infection among children ages 12 to 17 years by combining data from rounds 14 to 16 (to increase statistical power). A child was considered to have been vaccinated (single dose) 14 days after administration of the vaccine and to have been unvaccinated otherwise; similarly, a child was considered to have had two doses of vaccine 14 days after the second dose. We estimated vaccine effectiveness against infection as 1 - odds ratio (OR), where OR was estimated from a logistic regression model comparing swab positivity among vaccinated and unvaccinated children: adjustments were made for round and age, then additionally sex and IMD, region and ethnicity. Using data from round 16, we also estimated adjusted odds of infection by comparing swab positivity among adults who had received three doses of vaccine with those who had received two doses, allowing for a lag period of 14 days post-vaccination as above. Adjustments were made for age and sex, and, additionally, IMD, region and ethnicity.

## Data availability

Access to REACT-1 data is restricted due to ethical and security considerations. Summary statistics and descriptive tables from the current REACT-1 study are available in the Supplementary Information. Additional summary statistics and results from the REACT-1 programme are also available at https://www.imperial.ac.uk/medicine/research-and-impact/groups/react-study/real-time-assessment-of-community-transmission-findings/ and https://github.com/mrc-ide/reactidd/tree/master/inst/extdata REACT-1 Study Materials are available for each round at https://www.imperial.ac.uk/medicine/research-and-impact/groups/react-study/react-1-study-materials/

Sequence read data are available without restriction from the European Nucleotide Archive at https://www.ebi.ac.uk/ena/browser/view/PRJEB37886, and consensus genome sequences are available from the Global initiative on sharing all influenza data at https://www.gisaid.org.

## Data Availability

Access to REACT-1 data is restricted due to ethical and security considerations. Summary statistics and descriptive tables from the current REACT-1 study are available in the Supplementary Information. Additional summary statistics and results from the REACT-1 programme are also available at https://www.imperial.ac.uk/medicine/research-and-impact/groups/react-study/real-time-assessment-of-community-transmission-findings/ and https://github.com/mrc-ide/reactidd/tree/master/inst/extdata REACT-1 Study Materials are available for each round at https://www.imperial.ac.uk/medicine/research-and-impact/groups/react-study/react-1-study-materials/
Sequence read data are available without restriction from the European Nucleotide Archive at https://www.ebi.ac.uk/ena/browser/view/PRJEB37886, and consensus genome sequences are available from the Global initiative on sharing all influenza data at https://www.gisaid.org.

## Ethics

We obtained research ethics approval from the South Central-Berkshire B Research Ethics Committee (IRAS ID: 283787).

## Public involvement

A Public Advisory Panel provides input into the design, conduct, and dissemination of the REACT research program.

## Contributors

PE and CAD are corresponding authors. PE, MC-H and CAD conceived the study and the analytical plan. MC-H, OE, BB, HWang, DH, JJ, DT and CEW performed the statistical analyses. HWang, OE, DH, BB, and MW curated the data. JE, CA, PJD, DA, WB, GT, GC, HW, AD provided study oversight and results interpretation. AJP and AJT generated the sequencing data. AD and PE obtained funding. All authors revised the manuscript for important intellectual content and approved the submission of the manuscript. PE, MC-H, CAD had full access to the data and take responsibility for the integrity of the data and the accuracy of the data analysis and for the decision to submit for publication.

## Acknowledgements

PE is Director of the Medical Research Council (MRC) Centre for Environment and Health (MR/L01341X/1, MR/S019669/1). PE acknowledges support from Health Data Research UK (HDR UK); the NIHR Imperial Biomedical Research Centre; NIHR Health Protection Research Units in Chemical and Radiation Threats and Hazards, and Environmental Exposures and Health; the British Heart Foundation Centre for Research Excellence at Imperial College London (RE/18/4/34215); and the UK Dementia Research Institute at Imperial College London (MC_PC_17114). AJP acknowledges the support of the Biotechnology and Biological Sciences Research Council (BB/R012504/1). HW acknowledges support from an NIHR Senior Investigator Award, the Wellcome Trust (205456/Z/16/Z), and the NIHR Applied Research Collaboration (ARC) North West London. MC-H and BB acknowledge support from Cancer Research UK, Population Research Committee Project grant ‘Mechanomics’ (grant No 22184 to MC-H). MC-H and MW acknowledge support from the H2020-EXPANSE project (Horizon 2020 grant No 874627). CAD acknowledges support from the MRC Centre for Global Infectious Disease Analysis and National Institute for Health Research (NIHR) Health Protection Research Unit (HPRU) in Emerging and Zoonotic Infections. GC is supported by an NIHR Professorship.

We thank key collaborators on this work – Ipsos MORI: Kelly Beaver, Sam Clemens, Gary Welch, Nicholas Gilby, Kelly Ward, Galini Pantelidou and Kevin Pickering; Institute of Global Health Innovation at Imperial College London: Gianluca Fontana, Justine Alford; School of Public Health, Imperial College London: Eric Johnson, Rob Elliott, Graham Blakoe; Quadram Institute, Norwich, UK: Nabil-Fareed Alikhan; North West London Pathology and Public Health England (now UKHSA) for help in calibration of the laboratory analyses; Patient Experience Research Centre at Imperial College London and the REACT Public Advisory Panel; NHS Digital for access to the NHS register; the Department of Health and Social Care for logistic support.

## Supplementary Materials

**Supplementary Table 1.** Unweighted and weighted prevalence of swab-positivity from REACT-1 across rounds 1 to 16.

| Round | Tested swabs | Positive swabs | Unweighted prevalence (95% CI) | Weighted prevalence (95% CI) | First sample | Last sample |
| --- | --- | --- | --- | --- | --- | --- |
| 1 | 120,620 | 159 | 0.13% (0.11%, 0.15%) | 0.16% (0.13%, 0.19%) | 01/05/20 | 01/06/20 |
| 2 | 159,199 | 123 | 0.08% (0.07%, 0.09%) | 0.09% (0.07%, 0.11%) | 19/06/20 | 07/07/20 |
| 3 | 162,821 | 54 | 0.03% (0.03%, 0.04%) | 0.04% (0.03%, 0.05%) | 24/07/20 | 11/08/20 |
| 4 | 154,325 | 137 | 0.09% (0.08%, 0.11%) | 0.13% (0.01%, 0.15%) | 20/08/20 | 08/09/20 |
| 5 | 174,949 | 824 | 0.47% (0.44%, 0.50%) | 0.60% (0.55%, 0.71%) | 18/09/20 | 05/10/20 |
| 6 | 160,175 | 1,732 | 1.08% (1.03%, 1.13%) | 1.30% (1.21%, 1.39%) | 16/10/20 | 02/11/20 |
| 7 | 168,181 | 1,299 | 0.77% (0.73%, 0.82%) | 0.94% (0.87%, 1.01%) | 13/11/20 | 03/12/20 |
| 8 | 167,642 | 2,282 | 1.36% (1.31%, 1.42%) | 1.57% (1.49%, 1.66%) | 06/01/21 | 22/01/21 |
| 9 | 165,456 | 689 | 0.42% (0.39%, 0.45%) | 0.49% (0.44%, 0.55%) | 04/02/21 | 23/02/21 |
| 10 | 140,844 | 227 | 0.16% (0.14%, 0.18%) | 0.20% (0.17%, 0.23%) | 11/03/21 | 30/03/21 |
| 11 | 127,408 | 115 | 0.09% (0.07%, 0.11%) | 0.10% (0.08%, 0.13%) | 15/04/21 | 03/05/21 |
| 12* | 108,911 | 135 | 0.12% (0.10%, 0.15%) | 0.15% (0.12%, 0.18%) | 20/05/21 | 07/06/21 |
| 13 | 98,233 | 527 | 0.54% (0.49%, 0.58%) | 0.63% (0.57%, 0.69%) | 24/06/21 | 12/07/21 |
| 14** | 100,527 | 764 | 0.76% (0.71%, 0.82%) | 0.83% (0.76%, 0.89%) | 09/09/21 | 27/09/21 |
| 15*** | 100,112 | 1,399 | 1.40% (1.33%, 1.47%) | 1.57% (1.48%, 1.66%) | 19/10/21 | 05/11/21 |
| 16**** | 97,089 | 1,192 | 1.23% (1.16%, 1.30%) | 1.41% (1.33%, 1.51%) | 23/11/21 | 14/12/21 |

\* Sampling strategy changed for round 12 and subsequent rounds. Therefore unweighted prevalence is not directly comparable with previous rounds
\*\* Including N=509 samples from 28-30 September. Sample handling changed in round 14. Therefore prevalence is not directly comparable with previous rounds
\*\*\* Including N=93 samples (all negatives) from 6-8 November
\*\*\*\* Including N=661 samples (including 12 positives ) from 15-17 December

**Supplementary Table 2.** Proportion of each lineage detected in 650 positive samples with at least 50% genome coverage from round 16. Results are based on 1,048 samples collected up to and including 11 December 2021.

| Lineage | N (650) | Proportion |
| --- | --- | --- |
| AY.111 | 5 | 0.008 ( 0.003 , 0.018 ) |
| AY.116 | 5 | 0.008 ( 0.003 , 0.018 ) |
| AY.120 | 8 | 0.012 ( 0.006 , 0.024 ) |
| AY.121 | 1 | 0.002 ( 0.000 , 0.009 ) |
| AY.122 | 6 | 0.009 ( 0.004 , 0.020 ) |
| AY.122.1 | 5 | 0.008 ( 0.003 , 0.018 ) |
| AY.125 | 2 | 0.003 ( 0.001 , 0.011 ) |
| AY.127 | 1 | 0.002 ( 0.000 , 0.009 ) |
| AY.21 | 1 | 0.002 ( 0.000 , 0.009 ) |
| AY.25 | 2 | 0.003 ( 0.001 , 0.011 ) |
| AY.29.1 | 1 | 0.002 ( 0.000 , 0.009 ) |
| AY.34 | 5 | 0.008 ( 0.003 , 0.018 ) |
| AY.36 | 3 | 0.005 ( 0.002 , 0.013 ) |
| AY.39 | 1 | 0.002 ( 0.000 , 0.009 ) |
| AY.4 | 270 | 0.415 ( 0.378 , 0.454 ) |
| AY.4.2 | 86 | 0.132 ( 0.108 , 0.161 ) |
| AY.4.2.1 | 20 | 0.031 ( 0.020 , 0.047 ) |
| AY.4.2.2 | 9 | 0.014 ( 0.007 , 0.026 ) |
| AY.4.2.3 | 2 | 0.003 ( 0.001 , 0.011 ) |
| AY.42 | 4 | 0.006 ( 0.002 , 0.016 ) |
| AY.43 | 91 | 0.140 ( 0.115 , 0.169 ) |
| AY.44 | 1 | 0.002 ( 0.000 , 0.009 ) |
| AY.46 | 5 | 0.008 ( 0.003 , 0.018 ) |
| AY.46.5 | 3 | 0.005 ( 0.002 , 0.013 ) |
| AY.5 | 20 | 0.031 ( 0.020 , 0.047 ) |
| AY.5.3 | 3 | 0.005 ( 0.002 , 0.013 ) |
| AY.59 | 1 | 0.002 ( 0.000 , 0.009 ) |
| AY.6 | 6 | 0.009 ( 0.004 , 0.020 ) |
| AY.75 | 1 | 0.002 ( 0.000 , 0.009 ) |
| AY.87 | 1 | 0.002 ( 0.000 , 0.009 ) |
| AY.9 | 2 | 0.003 ( 0.001 , 0.011 ) |
| AY.9.1 | 3 | 0.005 ( 0.002 , 0.013 ) |
| AY.9.2 | 1 | 0.002 ( 0.000 , 0.009 ) |
| AY.90 | 1 | 0.002 ( 0.000 , 0.009 ) |
| AY.98 | 27 | 0.042 ( 0.029 , 0.060 ) |
| AY.98.1 | 2 | 0.003 ( 0.001 , 0.011 ) |
| B.1.617.2 | 34 | 0.052 ( 0.038 , 0.072 ) |
| BA.1 | 11 | 0.017 ( 0.009 , 0.030 ) |

\* Note lineage data are only available up to 11 December

**Supplementary Table 3.** Numbers of RT-PCR tests, positive tests, and Delta and Omicron infections identified by sequencing with 50% genome coverage (all Delta or Omicron) by day of swabbing

| Day of swab | Total tests | Positive tests | Positive tests with at least 50% coverage | Delta* | Omicron* |
| --- | --- | --- | --- | --- | --- |
| 2021-11-23 | 2,579 | 35 | 18 | 18 | 0 |
| 2021-11-24 | 5,292 | 62 | 38 | 38 | 0 |
| 2021-11-25 | 6,821 | 85 | 53 | 53 | 0 |
| 2021-11-26 | 6,013 | 68 | 43 | 43 | 0 |
| 2021-11-27 | 3,549 | 56 | 39 | 39 | 0 |
| 2021-11-28 | 2,447 | 25 | 17 | 17 | 0 |
| 2021-11-29 | 12,536 | 113 | 78 | 78 | 0 |
| 2021-11-30 | 10,794 | 123 | 84 | 84 | 0 |
| 2021-12-01 | 6,421 | 76 | 39 | 39 | 0 |
| 2021-12-02 | 5,868 | 70 | 41 | 41 | 0 |
| 2021-12-03 | 4,961 | 54 | 33 | 32 | 1 |
| 2021-12-04 | 2,488 | 27 | 16 | 16 | 0 |
| 2021-12-05 | 1,383 | 25 | 17 | 17 | 0 |
| 2021-12-06 | 5,279 | 55 | 42 | 42 | 0 |
| 2021-12-07 | 3,846 | 55 | 31 | 29 | 2 |
| 2021-12-08 | 3,227 | 43 | 25 | 22 | 3 |
| 2021-12-09 | 2,367 | 31 | 18 | 16 | 2 |
| 2021-12-10 | 2,776 | 27 | 12 | 10 | 2 |
| 2021-12-11 | 1,281 | 18 | 1 | 1 | 0 |
| 2021-12-12 | 796 | 17 | 0 | 0 | 0 |
| 2021-12-13 | 2,968 | 50 | 0 | 0 | 0 |
| 2021-12-14 | 1,544 | 33 | 0 | 0 | 0 |
| 2021-12-15 | 433 | 7 | 0 | 0 | 0 |
| 2021-12-16 | 150 | 2 | 0 | 0 | 0 |
| 2021-12-17 | 24 | 1 | 0 | 0 | 0 |
| Unknown | 1,135 | 21 | 5 | 4 | 1 |
| Total | 96,978 | 1,179 | 650 | 639 | 11 |

\* Note sequencing data only include samples up to 11 December 2021

**Supplementary Table 4.** Weighted prevalence of SARS-CoV2 swab-positivity in round 15 and round 16 by sex, age, region, urban *vs* rural environment, employment type, and ethnic group.

|  |  | Round 15 |  |  | Round 16 |  |  |
| --- | --- | --- | --- | --- | --- | --- | --- |
| Variable |  | Positive | Total | Weighted Prevalence | Positive | Total | Weighted Prevalence |
| Sex | Male | 679 | 44,887 | 1.66% (1.53%, 1.80%) | 564 | 43,938 | 1.49% (1.36%, 1.64%) |
|  | Female | 720 | 55,223 | 1.48% (1.36%, 1.60%) | 628 | 53,147 | 1.34% (1.23%, 1.46%) |
|  | Unknown | 0 | 2 |  | 0 | 4 |  |
| Age | 05-11 | 222 | 4,693 | 4.76% (4.16%, 5.44%) | 236 | 4,811 | 4.74% (4.15%, 5.40%) |
|  | 12-17 | 338 | 6,403 | 5.35% (4.78%, 5.99%) | 119 | 5,235 | 2.31% (1.91%, 2.80%) |
|  | 18-24 | 16 | 3,088 | 0.56% (0.33%, 0.95%) | 20 | 2,093 | 0.93% (0.57%, 1.51%) |
|  | 25-34 | 60 | 7,781 | 0.81% (0.62%, 1.07%) | 89 | 6,961 | 1.38% (1.10%, 1.74%) |
|  | 35-44 | 160 | 11,933 | 1.29% (1.09%, 1.51%) | 205 | 11,925 | 1.71% (1.48%, 1.98%) |
|  | 45-54 | 209 | 16,459 | 1.32% (1.14%, 1.52%) | 219 | 16,302 | 1.32% (1.15%, 1.52%) |
|  | 55-64 | 177 | 20,924 | 0.87% (0.74%, 1.01%) | 188 | 20,905 | 0.97% (0.84%, 1.13%) |
|  | 65-74 | 157 | 19,687 | 0.84% (0.72%, 0.99%) | 94 | 19,353 | 0.48% (0.39%, 0.59%) |
|  | 75+ | 60 | 9,144 | 0.63% (0.48%, 0.82%) | 22 | 9,504 | 0.21% (0.13%, 0.32%) |
| Region | South East | 261 | 17,637 | 1.62% (1.41%, 1.84%) | 228 | 16,908 | 1.54% (1.33%, 1.77%) |
|  | North East | 64 | 4,479 | 1.79% (1.36%, 2.34%) | 43 | 4,289 | 1.00% (0.70%, 1.42%) |
|  | North West | 179 | 12,079 | 1.65% (1.40%, 1.95%) | 107 | 11,352 | 1.08% (0.86%, 1.35%) |
|  | Yorkshire and The Humber | 136 | 9,720 | 1.43% (1.18%, 1.73%) | 96 | 9,261 | 1.32% (1.05%, 1.66%) |
|  | East Midlands | 110 | 8,647 | 1.47% (1.20%, 1.80%) | 125 | 8,584 | 1.73% (1.42%, 2.11%) |
|  | West Midlands | 135 | 9,943 | 1.59% (1.32%, 1.91%) | 89 | 9,715 | 1.03% (0.81%, 1.31%) |
|  | East of England | 158 | 11,560 | 1.57% (1.32%, 1.86%) | 121 | 11,437 | 1.22% (0.99%, 1.49%) |
|  | London | 160 | 14,847 | 1.23% (1.03%, 1.47%) | 239 | 14,819 | 1.84% (1.59%, 2.12%) |
|  | South West | 196 | 11,200 | 1.97% (1.69%, 2.29%) | 144 | 10,724 | 1.57% (1.30%, 1.90%) |
| Living in urban area | Yes | 1,091 | 78,469 | 1.54% (1.44%, 1.65%) | 956 | 75,982 | 1.43% (1.33%, 1.54%) |
|  | No | 306 | 21,477 | 1.69% (1.50%, 1.90%) | 233 | 20,951 | 1.35% (1.17%, 1.55%) |
|  | Unknown | 2 | 166 | 0.79% (0.19%, 3.23%) | 3 | 156 | 2.25% (0.70%, 6.98%) |
| Employment type | Health care or care home worker | 92 | 7,649 | 1.53% (1.23%, 1.91%) | 92 | 7,694 | 1.41% (1.13%, 1.76%) |
|  | Other essential/key worker | 237 | 14,065 | 1.87% (1.63%, 2.15%) | 247 | 13,427 | 1.96% (1.70%, 2.25%) |
|  | Other worker | 476 | 38,433 | 1.38% (1.25%, 1.53%) | 525 | 38,338 | 1.52% (1.38%, 1.68%) |
|  | Not full-time, part-time, or self-employed | 518 | 37,892 | 1.53% (1.39%, 1.68%) | 303 | 35,679 | 1.06% (0.92%, 1.21%) |
|  | Unknown | 76 | 2,073 | 3.50% (2.76%, 4.44%) | 25 | 1,951 | 1.19% (0.77%, 1.83%) |
| Ethnic group | White | 1,202 | 87,741 | 1.53% (1.44%, 1.63%) | 1,003 | 85,420 | 1.35% (1.26%, 1.45%) |
|  | Asian | 91 | 5,243 | 2.00% (1.58%, 2.53%) | 66 | 4,874 | 1.64% (1.24%, 2.17%) |
|  | Black | 34 | 1,919 | 1.66% (1.16%, 2.37%) | 38 | 1,833 | 2.10% (1.48%, 2.98%) |
|  | Mixed | 38 | 1,781 | 2.24% (1.59%, 3.14%) | 31 | 1,600 | 2.03% (1.40%, 2.94%) |
|  | Other | 12 | 1,074 | 1.22% (0.67%, 2.22%) | 16 | 984 | 1.81% (1.08%, 3.01%) |
|  | Unknown | 22 | 2,354 | 1.09% (0.69%, 1.71%) | 38 | 2,378 | 1.96% (1.40%, 2.74%) |

**Supplementary Table 5.** Estimates of vaccine effectiveness against infection for children aged 12 to 17 years in round 14 to round 16. Estimates are based on a logistic model of swab positivity in (i) children having received a single vaccine dose and (ii) children having received one or two vaccine doses compared to unvaccinated children. Results are all adjusted for round and and additionally adjusted for sex, and for Index of Multiple Deprivation (IMD), region, and ethnicity.

|  |  | Test negatives | Test positives | Vaccine Effectiveness |
| --- | --- | --- | --- | --- |
| Unvaccinated |  | 7,608 | 265 | - |
| 1 dose | Round, Age | 5,391 | 91 | 53.1% (39.5%, 63.7%) |
|  | Round, Age, Sex |  |  | 53.5% (38.7%, 64.8%) |
|  | Round, Age, Sex, IMD, Region, Ethnicity |  |  | 55.7% (41.3%, 66.6%) |
| 1 or 2 doses | Round, Age | 5,943 | 93 | 56.4% (43.9%, 66.2%) |
|  | Round, Age, Sex |  |  | 55.8% (41.6%, 66.6%) |
|  | Round, Age, Sex, IMD, Region, Ethnicity |  |  | 57.9% (44.1%, 68.3%) |

**Supplementary Table 6.** Estimates of the effect of a third vaccine dose on the risk of infection among participants of REACT-1 in round 16 compared to those who had received two vaccine doses. Results are presented for adults aged 18 years and over in round 16. Odds Ratios (ORs) are shown unadjusted, adjusted for age and sex, and additionally adjusted for Index of Multiple Deprivation (IMD), region, and ethnicity.

|  |  | Test negatives | Test positives | OR (95% CI) |
| --- | --- | --- | --- | --- |
| 2 Doses* |  | 40,603 | 584 | - |
| 3 Doses** | Unadjusted | 33,311 | 110 | 0.23 (0.19, 0.28) |
|  | Age, Sex |  |  | 0.28 (0.22, 0.34) |
|  | Age, Sex, IMD, Region, Ethnicity |  |  | 0.27 (0.22, 0.34) |

\* Participants with two doses are defined as those having only received two vaccine doses more than 14 days prior to swabbing or a third dose administered less than 14 days prior to swabbing
\*\* Participants with three doses are those who received their third vaccine dose 14 days prior to swabbing.

**Supplementary Table 7.** Weighted prevalence of SARS-CoV-2 swab-positivity in round 15 and round 16 by household size, number of children in the household, contact with a COVID-19 case, symptom status and neighbourhood deprivation.

|  |  | Round 15 |  |  | Round 16 |  |  |
| --- | --- | --- | --- | --- | --- | --- | --- |
| Variable |  | Positive | Total | Weighted Prevalence | Positive | Total | Weighted Prevalence |
| Household size | 1 | 132 | 16,997 | 0.78% (0.65%, 0.94%) | 122 | 16,657 | 0.88% (0.72%, 1.09%) |
|  | 2 | 318 | 39,624 | 0.83% (0.74%, 0.94%) | 287 | 39,443 | 0.77% (0.68%, 0.88%) |
|  | 3 | 283 | 17,579 | 1.76% (1.54%, 2.00%) | 226 | 16,626 | 1.40% (1.22%, 1.62%) |
|  | 4 | 421 | 17,774 | 2.48% (2.23%, 2.74%) | 362 | 17,026 | 2.29% (2.04%, 2.57%) |
|  | 5 | 178 | 5,752 | 3.12% (2.66%, 3.66%) | 137 | 5,206 | 2.73% (2.25%, 3.32%) |
|  | 6+ | 67 | 2,386 | 2.95% (2.27%, 3.82%) | 58 | 2,131 | 2.65% (2.00%, 3.50%) |
| Number of children in the household | 0 | 491 | 66,590 | 0.74% (0.67%, 0.81%) | 486 | 65,184 | 0.85% (0.76%, 0.95%) |
|  | 1+ | 644 | 27,239 | 2.62% (2.41%, 2.85%) | 621 | 26,709 | 2.43% (2.23%, 2.65%) |
|  | Unknown | 264 | 6,283 | 4.22% (3.72%, 4.78%) | 85 | 5,196 | 1.69% (1.35%, 2.12%) |
| COVID case contact | No | 595 | 80,225 | 0.83% (0.75%, 0.90%) | 529 | 75,990 | 0.81% (0.73%, 0.89%) |
|  | Yes, contact with a confirmed/tested COVID-19 case | 554 | 6,509 | 9.13% (8.35%, 9.96%) | 463 | 6,048 | 8.00% (7.25%, 8.82%) |
|  | Yes, contact with a suspected COVID-19 case | 67 | 1,448 | 5.04% (3.88%, 6.51%) | 51 | 1,523 | 3.29% (2.40%, 4.51%) |
|  | Unknown | 183 | 11,930 | 1.63% (1.39%, 1.91%) | 149 | 13,528 | 1.25% (1.04%, 1.49%) |
| Symptom status | Classic COVID symptoms* | 655 | 8,351 | 7.84% (7.22%, 8.51%) | 525 | 7,818 | 6.96% (6.32%, 7.67%) |
|  | Other symptoms | 208 | 15,634 | 1.47% (1.27%, 1.70%) | 211 | 15,377 | 1.52% (1.31%, 1.76%) |
|  | No symptoms | 357 | 64,231 | 0.67% (0.60%, 0.75%) | 308 | 60,413 | 0.62% (0.55%, 0.70%) |
|  | Unknown | 179 | 11,896 | 1.60% (1.36%, 1.88%) | 148 | 13,481 | 1.24% (1.03%, 1.49%) |
| Deprivation | 1 Most deprived | 174 | 11,556 | 1.70% (1.45%, 1.99%) | 138 | 11,127 | 1.47% (1.21%, 1.78%) |
|  | 2 | 215 | 16,850 | 1.41% (1.22%, 1.63%) | 245 | 16,263 | 1.61% (1.40%, 1.84%) |
|  | 3 | 283 | 21,096 | 1.51% (1.33%, 1.72%) | 252 | 20,455 | 1.39% (1.21%, 1.58%) |
|  | 4 | 327 | 24,130 | 1.55% (1.38%, 1.74%) | 255 | 23,307 | 1.26% (1.10%, 1.44%) |
|  | 5 Least deprived | 400 | 26,480 | 1.67% (1.51%, 1.86%) | 302 | 25,937 | 1.37% (1.21%, 1.54%) |

\* Classic COVID symptoms: loss or change of sense of smell or taste, fever, new persistent cough

**Supplementary Table 8.** Multivariable logistic regression for round 15 and round 16. Results are presented as Odds Ratios (95% confidence interval) adjusted for age and sex and additionally for all other variables (mutually adjusted OR).

| Variable |  | Adjusted for age and sex | Mutually adjusted* |
| --- | --- | --- | --- |
| Gender | Male | Ref | Ref |
|  | Female | 0.90 (0.80, 1.01) | 0.92 (0.82, 1.03) |
| Age | 05-11 | 2.93 (2.42, 3.54) | 2.65 (2.18, 3.23) |
|  | 12-17 | 1.32 (1.05, 1.66) | 1.68 (1.22, 2.32) |
|  | 18-24 | 0.55 (0.35, 0.88) | 0.64 (0.40, 1.03) |
|  | 25-34 | 0.74 (0.58, 0.95) | 0.85 (0.65, 1.10) |
|  | 35-44 | Ref | Ref |
|  | 45-54 | 0.78 (0.64, 0.94) | 0.84 (0.70, 1.03) |
|  | 55-64 | 0.52 (0.42, 0.63) | 0.68 (0.54, 0.85) |
|  | 65-74 | 0.28 (0.22, 0.35) | 0.39 (0.29, 0.52) |
|  | 75+ | 0.13 (0.08, 0.20) | 0.19 (0.12, 0.30) |
| Region | South East | Ref | Ref |
|  | North East | 0.77 (0.56, 1.07) | 0.76 (0.55, 1.06) |
|  | North West | 0.73 (0.58, 0.93) | 0.73 (0.58, 0.93) |
|  | Yorkshire and The Humber | 0.78 (0.61, 1.00) | 0.78 (0.61, 0.99) |
|  | East Midlands | 1.11 (0.89, 1.39) | 1.11 (0.89, 1.39) |
|  | West Midlands | 0.68 (0.53, 0.87) | 0.67 (0.52, 0.87) |
|  | East of England | 0.78 (0.63, 0.98) | 0.78 (0.62, 0.97) |
|  | London | 1.05 (0.87, 1.26) | 1.04 (0.86, 1.26) |
|  | South West | 1.04 (0.84, 1.29) | 1.02 (0.83, 1.27) |
| Employment type | Health care or care home worker | 0.74 (0.59, 0.93) | 0.75 (0.59, 0.94) |
|  | Other essential/key worker | 1.18 (1.01, 1.38) | 1.18 (1.01, 1.38) |
|  | Other worker | Ref | Ref |
|  | Not full-time, part-time, or self-employed | 0.93 (0.79, 1.09) | 0.95 (0.81, 1.12) |
|  | Unknown | 0.63 (0.41, 0.96) | 0.64 (0.42, 0.98) |
| Ethnic group | White | Ref | Ref |
|  | Asian | 0.78 (0.61, 1.01) | 0.74 (0.57, 0.95) |
|  | Black | 1.24 (0.89, 1.73) | 1.16 (0.83, 1.64) |
|  | Mixed | 0.92 (0.64, 1.33) | 0.88 (0.61, 1.27) |
|  | Other | 1.05 (0.64, 1.73) | 0.96 (0.58, 1.60) |
|  | Unknown | 1.44 (1.04, 2.01) | 1.39 (1.00, 1.94) |
| Household size | 1-2 | Ref | Ref |
|  | 3-5 | 1.30 (1.12, 1.51) | 1.13 (0.94, 1.35) |
|  | 6+ | 1.63 (1.21, 2.20) | 1.39 (1.01, 1.91) |
| Number of children in the household | 0 | Ref | Ref |
|  | 1+ | 1.47 (1.25, 1.74) | 1.37 (1.12, 1.67) |
|  | Unknown | 0.99 (0.69, 1.41) | 0.95 (0.66, 1.38) |
| Living in urban area | Yes | Ref | Ref |
|  | No | 0.99 (0.86, 1.14) | 1.00 (0.85, 1.16) |
|  | Unknown | 1.54 (0.49, 4.87) | 1.61 (0.51, 5.09) |
| Deprivation | 1 Most deprived | 0.95 (0.78, 1.17) | 1.06 (0.86, 1.31) |
|  | 2 | 1.23 (1.04, 1.46) | 1.28 (1.07, 1.52) |
|  | 3 | 1.05 (0.89, 1.24) | 1.08 (0.91, 1.28) |
|  | 4 | 0.96 (0.81, 1.14) | 0.99 (0.83, 1.17) |
|  | 5 Least deprived | Ref | Ref |

\* Odds ratios are mutually adjusted for all variables shown.

**Supplementary Figure 1.**
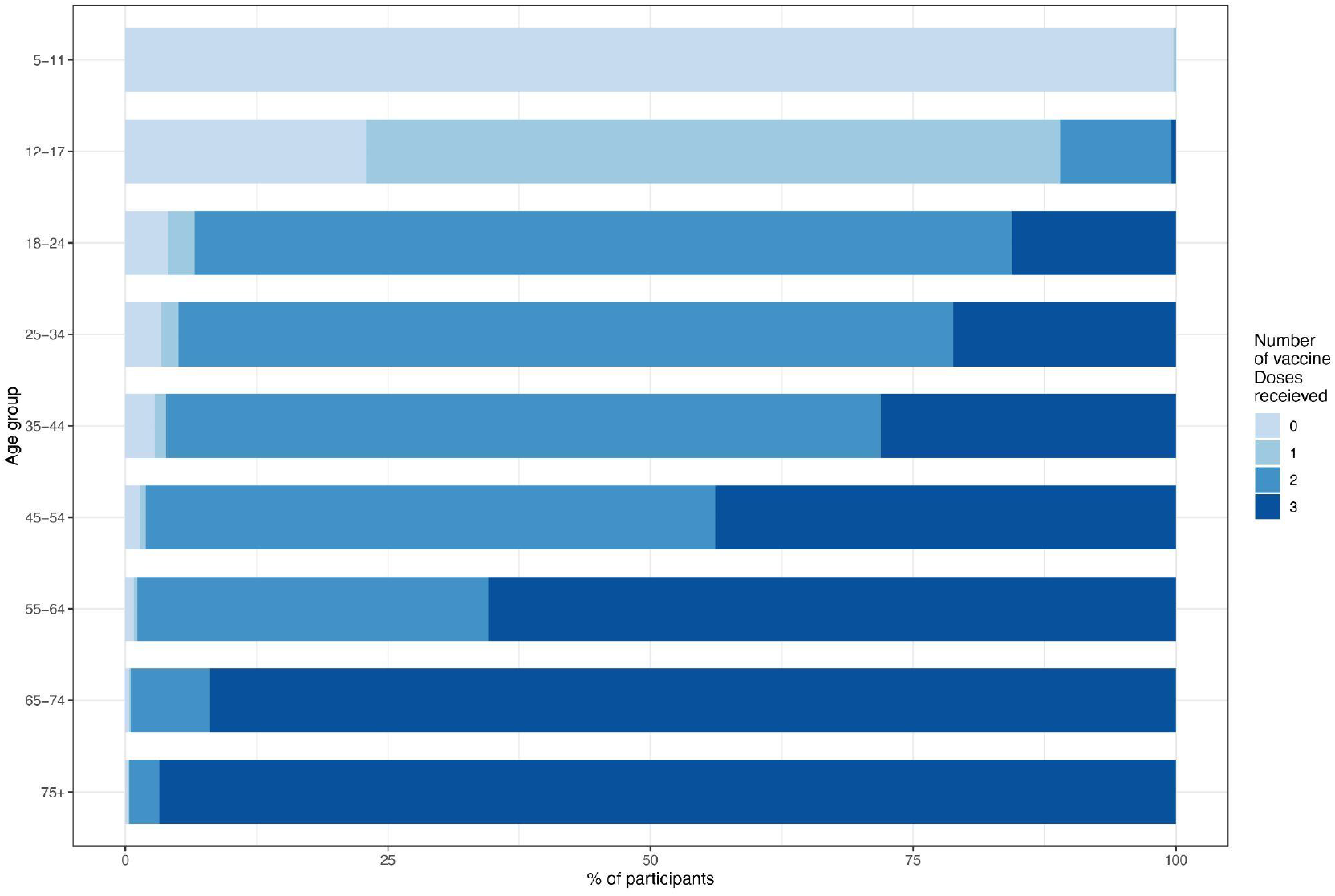
Proportion of unvaccinated (pale blue) participants and participants having received one, two, or three vaccine doses by age in round 16. Results are based on linked vaccination data in consenting participants.

